# Autonomic Modulation with Mindfulness-Based Stress Reduction in Chronic Kidney Disease: A Randomized Controlled Trial

**DOI:** 10.1101/2024.04.17.24306000

**Authors:** Jinhee Jeong, Yingtian Hu, Matias Zanuzzi, Dana DaCosta, Sabrina Li, Jeanie Park

## Abstract

**Background:** Chronic kidney disease (CKD) is characterized by overactivation of the sympathetic nervous system (SNS) that leads to increased cardiovascular disease risk. Despite the deleterious consequences of SNS overactivity, there are very few therapeutic options available to combat sympathetic overactivity.

**Aim:** To evaluate the effects of Mindfulness-Based Stress Reduction (MBSR) on SNS activity in CKD patients.

**Method:** Participants with CKD stages III-IV were randomized to an 8-week MBSR program or Health Education Program (HEP; a structurally parallel, active control group). Primary outcomes were direct intraneural measures of SNS activity directed to muscle (MSNA) via microneurography at rest and during stress maneuvers.

**Results:** 28 participants (63 ±9 years; 86% males) completed the intervention with 16 in MBSR and 12 in HEP. There was a significant Group (MBSR vs. HEP) by Time (baseline vs. post-intervention) interaction in the change in MSNA reactivity to mental stress (p=0.026), with a significant reduction in the mean change in MSNA over 3 minutes of mental arithmetic at post-intervention (10.6 ± 7.1 to 5.0 ± 5.7 bursts/min, p<0.001), while no change was observed within the HEP group (p=0.773).

**Conclusions:** In this randomized controlled trial, patients with CKD had an amelioration of sympathetic reactivity during mental stress following 8-weeks of MBSR but not after HEP. Our findings demonstrate that mindfulness training is feasible and may have clinically beneficial effects on autonomic function in CKD.

## Introduction

Chronic kidney disease (CKD) afflicts more than 35 million individuals in the United States and is associated with a significantly elevated risk of cardiovascular (CV) disease and mortality by up to 5-15 fold ^1, 2^. A primary contributor to heightened CV risk among these patients is the persistent elevation of sympathetic nervous system (SNS) activity ^3–5^. Even modest declines in kidney function lead to substantial increases in both resting SNS activity as well as SNS reactivity that are independently associated with adverse CV events ^3, 5^. The use of sympatholytic medications in CKD is clinically limited ^6, 7^ due to adverse side effects ^8–10^ and inferior efficacy in CV risk reduction compared to alternative antihypertensive agents ^8, 11, 12^. Therefore, there is a critical need to develop novel therapeutic approaches to safely and effectively counter SNS overactivity to improve clinical outcomes in CKD and other high-risk patient populations characterized by SNS overactivation.

Mindfulness meditation may be one such novel nonpharmacologic strategy to lower SNS activity in individuals with CKD. In particular, mindfulness-based stress reduction (MBSR) is an 8-week structured mindfulness meditation training program that has been extensively studied and has demonstrated beneficial effects on CV outcomes in patient populations ^13–15^. Specifically, some, although not all, studies have shown that MBSR lowers clinic and ambulatory blood pressure (BP) ^16–20^, and attenuates psychological ^21, 22^ and physiological reactivity to stress ^23–26^. Although the mechanisms underlying the cardioprotective mechanisms of MBSR remain unknown, indirect evidence suggests that MBSR may lower BP by modulating autonomic nervous system activity, particularly by lowering SNS activity ^27–29^. Earlier investigations using indirect measures of SNS activity have shown that meditation increases low frequency and high frequency heart rate variability ^30–32^ and lowers plasma norepinephrine levels ^33^. In addition, our group previously measured direct, intraneural recordings of muscle sympathetic nerve activity (MSNA) via the gold-standard microneurographic technique to demonstrate the first direct evidence that a single session of mindfulness meditation acutely reduces BP and MSNA in patients with CKD ^34^. However, there have been no prior studies testing the sustained effects of MBSR on SNS activity and reactivity using direct measures of SNS activity in humans. Therefore, we performed a randomized controlled trial to test the hypothesis that MBSR leads to sustained reductions in both resting MSNA and MSNA reactivity to stress in CKD.

## Methods

### Participants

Individuals with CKD stages III and IV (estimated glomerular filtration rate (eGFR) between 15-59 mL/min/1.73 m^2^), as defined by the race-free CKD-EPI equation ^35^, were recruited from Emory University clinics and the Atlanta Veterans Affairs (VA) Health Care System for participation in this study from September 2019 to Jun 2022. All participants had stable renal function (no greater than a decline of eGFR of 1cc/min/1.73m^2^ per month over the prior 3 months). Exclusion criteria included uncontrolled hypertension (BP>160/90mmHg), vascular disease, use of clonidine, clinical evidence of heart failure or active heart disease determined by history, electrocardiogram (ECG) or echocardiogram, ongoing illicit drug use, alcohol use > 2 drinks/day within the past 12 months, diabetic neuropathy, severe anemia (hemoglobin <10mg/dL), and pregnancy or plans to become pregnant.

### Study Design and Randomization

This was a single-site, parallel-group, randomized controlled clinical trial. Participants were allocated to MBSR group or a control Health-Enhancement Program (HEP) group in a 1:1 ratio using a computer-generated sequence stratified by stage of CKD (Stage IIIA, IIIB, and IV). While the nature of the interventions did not allow for masking of the MBSR and HEP instructors and participants, all investigators and assessors were masked to study identifiers or group assignment during data analysis. After obtaining written informed consent, baseline measurements and basic metabolic panel were obtained. All measurements were obtained in a quiet, temperate (21°C) environment, after abstaining from food, caffeine, smoking, and alcohol for at least 12 h, and exercise for at least 24 h. After baseline assessments, participants were randomly assigned to one of two interventions; MBSR and HEP. At the end of the trial, baseline measurements were repeated under identical conditions within 2 weeks following the completion of the final session. Participants were instructed not to change medication regimens, dietary habits, or physical activity levels for the duration of the study. This study was approved by the Emory University Institutional Review Board and the Atlanta VA Health Care System Research and Development Committee and registered with clinicaltrials.gov (NCT 04099992). All study procedures conformed to the standards set forth by the Declaration of Helsinki.

### Study Interventions

Both interventions followed detailed manuals and were administered to small groups (4-6 participants) for 2.5 hours, once per week for eight consecutive weeks. Weekly group sessions as well as a half-day retreat were administered using Zoom© video conferencing. In addition to weekly attendance, participants were assigned 45-minutes of homework 6 days per week.

### MBSR intervention

The MBSR intervention was comprised of 8 weeks of 2.5 h sessions held once per week. An additional one-day “retreat” was also required for participants to attend between weeks 7 and 8. A single, fully-certified teacher (M.D.) who had completed the University of Massachusetts MBSR professional training program with more than 15 years of mindfulness practice and teaching experience delivered the formalized program curriculum to participants.

The formal MBSR program focuses on mindfulness by increasing awareness of thoughts, feelings, and sensations from moment to moment, and to more skillfully respond (rather than automatically react) to stressors. Each of the eight weekly sessions includes psychoeducation about mindfulness and stress (including a discussion of habitual behavioral and physiological reactions to stress, and learning to more purposely respond rather than react); experiential mindfulness practice, and a discussion of participants’ experiences with mindfulness practice. Participants learn formal mindfulness practices (e.g., sitting meditation, yoga, body scan, walking meditation, body scan meditation) as well as more informal mindfulness activities such as awareness of breath, thoughts, or emotions, and mindfulness of daily life activities (e.g., eating, breathing, washing dishes). They were encouraged to practice mindfulness in their daily lives. Participants received MP4 downloads on an electronic tablet with guided mindfulness meditation practices, a home practice manual, and weekly handouts with each week’s formal and informal practice assignments. Participants were asked to perform daily home practice of 40-45 minutes of recorded practice and keep a written record of their practice times. A makeup class on a different day was available if a participant missed a class. The half-day retreat session between weeks 7 and 8 sought to further integrate acquired skills and involved guided meditation practice and reflection.

### HEP Intervention

An 8-week health enhancement program (HEP), a non-meditation intervention was administered as the gold-standard control intervention for MBSR ^36–38^. HEP is designed to provide a structurally parallel, active control intervention to MBSR within the same group-based environment, matching for group support, facilitator attention, intervention duration, time spent on at-home practice while omitting any components of mindfulness. The original HEP manual had been developed for use in a non-clinical sample as a control condition for MBSR ^38^. Specifically, HEP consists of music therapy, nutritional education, posture and balance movements, walking and stretching instructed by a health educator (a registered dietician) in a group setting for 8 weekly 2.5-hour sessions. Work with all practices with group discussion and exercises during an all-day match the all-day retreat in MBSR. Participants received MP4 downloads on an electronic tablet with recordings of health education topics, a home listening manual, and weekly handouts with each week’s listening assignments, and were asked to listen to these MP4 recordings daily for 40-45 minutes and log their daily adherence. HEP was administered by a clinician with health education experience.

### Outcomes and Measurements

The primary outcomes were sympathetic activity quantified as resting MSNA and MSNA reactivity to acute stressors. Secondary outcomes included resting BP and BP reactivity to acute stressors.

### Muscle Sympathetic Nerve Activity

Multiunit postganglionic MSNA was recorded directly from the peroneal nerve by microneurography, as previously described ^39^. Participants were placed in a supine position, and the leg was positioned for microneurography. A tungsten microelectrode (tip diameter 5–15 μm) (Bioengineering, University of Iowa) was inserted into the nerve, and a reference microelectrode was inserted subcutaneously 1–2 cm from the recording electrode. The signals were amplified (total gain: 50,000–100,000), filtered (700-2,000 Hz), rectified, and integrated (time constant 0.1 s) to obtain a mean voltage display of sympathetic nerve activity (Nerve Traffic Analyzer, model 662C-4, University of Iowa, Bioengineering) that was recorded by the LabChart 7 Program (PowerLab 16sp, ADInstruments). Continuous ECG was recorded simultaneously with the neurogram using a bioamp system. Beat-to-beat arterial BP was measured concomitantly using a noninvasive monitoring device that detects digital blood flow via finger cuffs and translates blood flow oscillations into continuous pulse pressure waveforms and beat-to-beat values of BP (Finometer, Finapres Medical Systems, Amsterdam, The Netherlands) ^40^. Absolute values of BP were internally calibrated using a concomitant upper arm BP reading and were calibrated at the start and every 15 min throughout the study. The tungsten microelectrode was manipulated to obtain a satisfactory nerve recording that met previously established criteria ^41^. After 10 min of rest, BP, ECG, and MSNA were recorded continuously for 10 min. After 10 minutes of rest, MSNA, beat-to-beat BP and ECG were continuously measured throughout the stress protocols that include mental arithmetic (3 min), handgrip exercise (3 min) and cold pressor test (CPT, 1min), separated by 15 min of rest and followed by 5 minutes of recovery. Detailed stress maneuver protocols are described below.

For MSNA data analysis, MSNA and ECG data were exported from the LabChart data acquisition system to WinCPRS (Absolute Aliens, Turku, Finland) for analysis. R-waves were detected and marked from the continuous ECG recording. MSNA bursts were automatically detected by the program using the following criteria: 3:1 burst-to-noise ratio within a 0.5-s search window, with an average latency in burst occurrence of 1.2–1.4 s from the previous R-wave. After automatic detection, the ECG and MSNA neurograms were visually inspected for accuracy of detection. MSNA was expressed as burst frequency (bursts/min) and burst incidence (bursts/100 heartbeats).

### Blood Pressure

Baseline BP was measured after 5 min of rest in a seated position with the arm supported at heart level using an appropriately sized cuff per American College of Cardiology/American Heart Association (ACC/AHA) guidelines ^11^ using an automated device (Omron, HEM-907XL, Omron Healthcare, Kyoto, Japan). Mean arterial blood pressures (MAP) were calculated as 2/3 diastolic BP (DBP) + 1/3 systolic BP (SBP).

### Intervention Satisfaction

Participants in both groups completed the 8-item Client Satisfaction Questionnaire (CSQ-8) to assess intervention satisfaction ^42^ at post-intervention.

### Stress Maneuvers

#### Mental Arithmetic

Participants were asked to serially subtract a one– or two-digit number from a three– or four-digit number and were urged to do so as quickly and accurately as possible for 3 minutes. An investigator used flash cards with a three– or four-digit number and were urged by two additional study team members in white coats to answer ‘faster’ and ‘get it right’. At the end of mental stress, participants were asked to rate their perceived stress during the mental math trial using a standard five-point scale, ranging from 0 (no stress) to 4 (severe stress). This test is shown to induce mental stress and increases MSNA ^43^ ^44^.

#### Static Handgrip (SHG)

Moderate intensity SHG was performed by squeezing the hand dynamometer at 30% maximal voluntary contraction in a sustained manner for 3 min. The participant was instructed to avoid inadvertent Valsalva and to maintain normal breathing patterns. Moderate SHG elicits an increase in MSNA ^45^ by activating muscle metaboreceptors and mechanoreceptors.

#### Cold Pressor Test (CPT)

The dominant hand was immersed up to the wrist in an ice water bath (4°C) for 1 min, which was followed by 3 min of recovery. Subjects were instructed to avoid breath holding and to stay as relaxed as possible. The participant was asked to rate the degree of pain during the maneuver on a five-point pain scale, ranging from 0 (no pain) to 4 (severe pain). CPT is a sympatho-excitatory maneuver that is known to evoke an increase in MSNA ^46^.

### Statistics

Based on our pilot data, the group difference in MSNA change after MBSR program (vs. a control program) was estimated to be –9.7 ± 5.4 bursts/min. 40 participants (20 per group) provided >90 % power at a significance level (α) of 0.05 for primary outcome (MSNA) after considering the expected 20% dropout rate.

Values are presented as mean ± standard deviation (SD) unless otherwise noted. Differences in participant characteristics between groups were determined using independent, 2-tailed, *t*-tests for continuous variables and chi-square tests for categorical variables. The primary analyses were based on the intention-to-treat approach. Repeated ANOVA tests with Group (MBSR vs. HEP) as a between factor and Time (baseline and post-intervention) as a within factor was used to analyze the change in MSNA reactivity during 3 minutes of mental arithmetic and SHG and 1 minute of CPT. When an ANOVA revealed a significant main effect or interaction, post hoc pairwise comparisons using the Sidak test were made. The within-group comparison from baseline to 8 weeks was done using 2-tailed, paired *t*-tests. In addition, a linear mixed model (LMM) analysis was conducted with Minutes, Group and Time as fixed factors and participants as a random factor to compare the group difference in slope-of-rise in the absolute MSNA change over 3 minutes of mental arithmetic and SHG. Adjustment with potential confounding factors, including the use of antihypertensive medications, was performed. An α <0.05 was considered statistically significant for all analyses. All analyses were performed using SPSS version 26.0 (IBM Corporation, Somers, NY) and R (version 4.0.2, R Foundation for Statistical Computing, Vienna, Austria).

## Results

### Participants and Intervention Adherence

A total of twenty-eight participants who completed the study intervention were included in this study (**Figure 1)**. No significant differences in baseline demographics, comorbid conditions, medication use or clinical characteristics were observed between the MBSR and HEP groups. Intervention adherence measured by attendance of intervention group sessions, retreats and time spent on total home practice was high in both groups and comparable. CSQ-8 score indicated that participants in both groups were generally satisfied with the intervention (**Table 1**).

**Figure 1.**
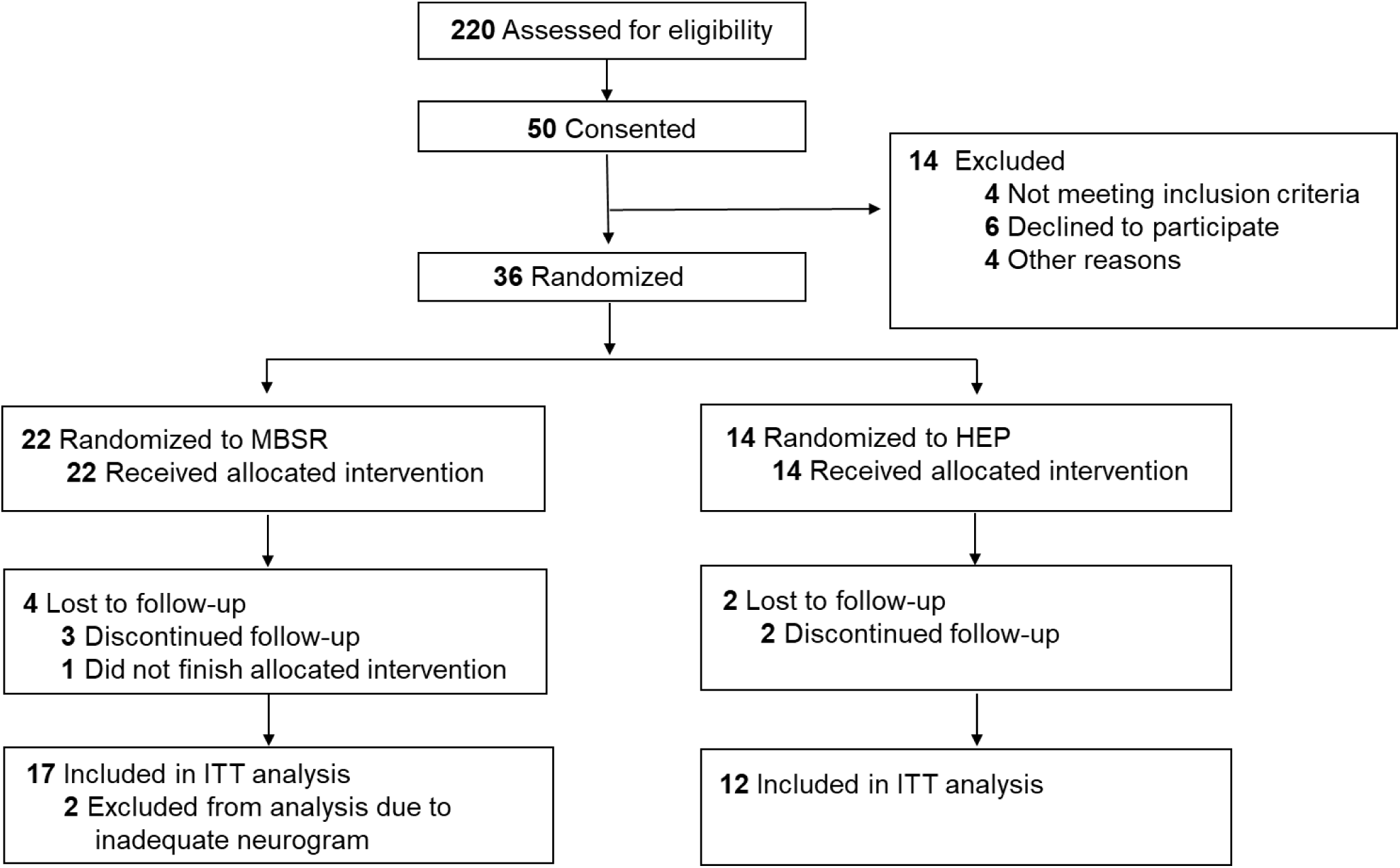
CONSORT diagram. The diagram depicts patient flow through the randomized clinical trial.

**Table 1.** Participant Characteristics and Intervention Adherence.

| Characteristics | MBSR (n=16) | HEP (n=12) |
| --- | --- | --- |
| **Age (yr)** | 64.1 ± 8.4 | 63.3 ± 10.3 |
| **Sex (male, %)** | 14 (88%) | 10 (83%) |
| **Race** |  |  |
| Black | 9 (56%) | 10 (83%) |
| White | 7 (44%) | 2 (17%) |
| **Height (cm)** | 175.5 ± 8.6 | 176.7 ± 10.7 |
| **Weight (kg)** | 101.4 ± 23.0 | 94.1 ± 16.9 |
| **BMI (kg/m<sup>2</sup>)** | 32.9 ± 6.4 | 30.1 ± 4.8 |
| **Smokers (n,%)** | 2 (13%) | 1 (8%) |
| **Hypertension (n,%)** | 13 (81%) | 11 (92%) |
| **Diabetes (n,%)** | 7 (44%) | 7 (58%) |
| **Antihypertensive medications (n,%)** |  |  |
| ACEI/ARB | 13 (81%) | 9 (75%) |
| CCB | 9 (56%) | 7 (58%) |
| Diuretics | 8 (50%) | 5 (42%) |
| β-blockers | 3 (19%) | 7 (58%) |
| α-blockers | 4 (25%) | 2 (17%) |
| Hydralazine | 2 (13%) | 0 (0%) |
| **Statin (n,%)** | 10 (63%) | 7 (58%) |
| **eGFR (ml·min<sup>-1</sup>·1.73 m<sup>-2</sup>)** | 39.9 ± 12.3 | 46.0 ± 7.4 |
| **Intervention Adherence and Satisfaction** |  |  |
| Class Attendance (%) | 95 ± 10 | 94 ± 11 |
| Home Practice (%) | 78 ± 23 | 85 ± 21 |
| Home Practice (hour) | 34 ± 21 | 43 ± 23 |
| Retreat Attendance | 14/16 | 8/12 |
| Satisfaction | 27 ± 4 (86 ± 13%) | 30 ± 2 (95 ± 5 %) |

Continuous values are expressed as means ± standard deviation.
**BMI:** Body Mass Index, **ACEI:** Angiotensin Converting Enzyme Inhibitors, **ARB:** Angiotensin Receptor Blocker **CCB:** Calcium Channel Blocker, **eGFR:** Estimated Glomerular Filtration Rate.

### Effect of MBSR on MSNA and Hemodynamics at Rest and During Stress

#### Resting Outcomes

Resting MSNA, BP and heart rate remained unchanged in both groups from pre-intervention baseline levels to post-intervention (**Table 2**).

**Table 2.** Effects of MBSR vs. HEP Intervention on Resting and Reactivity to Static Handgrip and Cold Pressor Test of Cardiovascular and ympathetic Parameters at in Patients with Chronic Kidney Disease.

|  | MBSR (n=16) |  |  | HEP (n=12) |  |  |  |
| --- | --- | --- | --- | --- | --- | --- | --- |
|  | Baseline | 8 weeks | Within-group P-value | Baseline | 8 weeks | Within-group P-Value | Group x Time P-value |
| **Resting** |  |  |  |  |  |  |  |
| SBP (mmHg) | 122 ± 11 | 118 ± 15 | 0.199 | 123 ± 12 | 121 ± 11 | 0.606 | 0.182 |
| DBP (mmHg) | 75 ± 8 | 70 ± 13 | 0.088 | 73 ± 12 | 72 ± 9 | 0.301 | 0.945 |
| MAP (mmHg) | 90 ± 8 | 86 ± 13 | 0.099 | 89 ± 11 | 88 ± 8 | 0.318 | 0.390 |
| HR (bpm) | 72 ± 9 | 69 ± 9 | 0.152 | 64 ± 7 | 66 ± 10 | 0.441 | 0.135 |
| MSNA (bursts/min) | 32.8 ± 7.6 | 33.8 ± 8.9 | 0.448 | 38.9 ± 9.4 | 36.2 ± 8.9 | 0.117 | 0.096 |
| MSNA (bursts/100HB) | 53.7 ± 15.3 | 55.8 ± 17.9 | 0.501 | 61.3 ± 12.3 | 59.3 ± 12.6 | 0.547 | 0.372 |
| **Reactivity to SHG** |  |  |  |  |  |  |  |
| Δ SBP (mmHg) | 10.5 ± 10.6 | 10.9 ± 13.2 | 0.922 | 10.8 ± 10.7 | 5.3 ± 8.3 | 0.205 | 0.286 |
| Δ DBP (mmHg) | 7 ± 7 | 9 ± 8 | 0.500 | 8 ± 6 | 6 ± 8 | 0.454 | 0.318 |
| Δ MAP (mmHg) | 10 ± 9 | 10 ± 10 | 0.976 | 10 ± 9 | 7 ± 6 | 0.318 | 0.416 |
| Δ HR (bpm) | 5 ± 4 | 6 ± 5 | 0.219 | 5 ± 6 | 5 ± 4 | 0.960 | 0.623 |
| Δ MSNA (bursts/min) | 14.3 ± 7.0 | 10.9 ± 6.4 | 0.139 | 9.3 ± 6.0 | 11.2 ± 9.1 | 0.484 | 0.142 |
| Δ MSNA (bursts/100HR) | 16.6 ± 11.4 | 9.9 ± 10.5 | 0.041 | 8.3 ± 7.9 | 11.8 ± 12.3 | 0.363 | 0.048 |
| **Reactivity to CPT** |  |  |  |  |  |  |  |
| Δ SBP (mmHg) | 7 ± 8 | 1 ± 17 | 0.109 | 16 ± 15 | 10 ± 11 | 0.242 | 0.943 |
| Δ DBP (mmHg) | 8 ± 4 | 8 ± 10 | 0.812 | 14 ± 13 | 13 ± 12 | 0.596 | 0.783 |
| Δ MAP (mmHg) | 9 ± 6 | 6 ± 12 | 0.392 | 17 ± 16 | 11 ± 10 | 0.318 | 0.833 |
| Δ HR (bpm) | 7 ± 5 | 6 ± 4 | 0.219 | 9 ± 9 | 10 ± 7 | 0.496 | 0.200 |
| Δ MSNA (bursts/min) | 13.5 ± 15.9 | 12.1 ± 5.8 | 0.563 | 18.6 ± 16.4 | 22.1 ± 8.5 | 0.446 | 0.341 |
| Δ MSNA (bursts/100HR) | 15.1 ± 22.1 | 13.4 ± 12.5 | 0.713 | 17.6 ± 12.9 | 21.8 ± 6.2 | 0.503 | 0.450 |

Values are expressed as mean ± standard deviation.
**SBP:** Systolic Blood Pressure, **DBP:** Brachial Diastolic Blood Pressure, **MAP:** Mean Arterial Blood Pressure, **HR:** Heart Rate, **MSNA:** Muscle Sympathetic Nervous Activity, **HB:** Heart Beats, **SHG:** Static Handgrip, **CPT:** Cold Pressor Test. Within-group comparisons over 8 weeks were tested by paired *t*-tests and between-group comparisons on the changes over 8 weeks (Group by Time) were tested by repeated ANOVA tests.

#### Reactivity to Mental Arithmetic

There was a significant Group (MBSR vs. HEP) by Time (baseline vs. post-intervention) interaction in the change in MSNA reactivity to mental arithmetic stress. The MBSR group had a significant reduction in the mean change in MSNA over 3 minutes of mental arithmetic at post-intervention (10.6 ± 7.1 to 5.0 ± 5.7 bursts/min; 13.3 ± 9.3 to 4.2 ± 8.5 bursts/100 heartbeats; p<0.001 for both), while no change was observed within the HEP group (9.7 ± 6.1 to 9.1 ± 5.6 bursts/min; 11.9 ± 8.7 to 14.3 ± 12.4 bursts/100 heartbeats; p=0.773 and 0.304 respectively) (**Figure 2 A-B**). MSNA reactivity was lower at the 1^st^, 2^nd^ and 3^rd^ minute of mental arithmetic within the MBSR group at post-intervention (p<0.05 for all), while no such change was observed within the HEP group (p>0.05 for all) (**Figure 3 A-D**). However, there was no significant change in the reactivity of BP and heart rate during mental arithmetic (**Figure 2 E-H**) In addition, LMM analyses revealed a significant decrease in the slope-of-rise of absolute MSNA changes during 3 minutes of mental arithmetic from baseline to post-intervention within the MBSR group, but not in the HEP group (effect sizes for the group difference (MBSR vs. HEP) in the slope change (baseline to post-intervention) of MSNA: –1.57 bursts/min per min; p=0.008 and –2.52 bursts/100 heartbeats per min; p=0.004, respectively). Results were similar when adjusted for antihypertensive medication usage. Perceived stress levels during mental arithmetic were comparable between baseline and post-intervention in both groups (2.2 ± 1.2 to 2.2 ± 0.9 in the MBSR and 2.5 ± 1.0 to 2.4 ± 1.3 in the HEP group, p=0.825 for Time by Group interaction).

**Figure 2.**
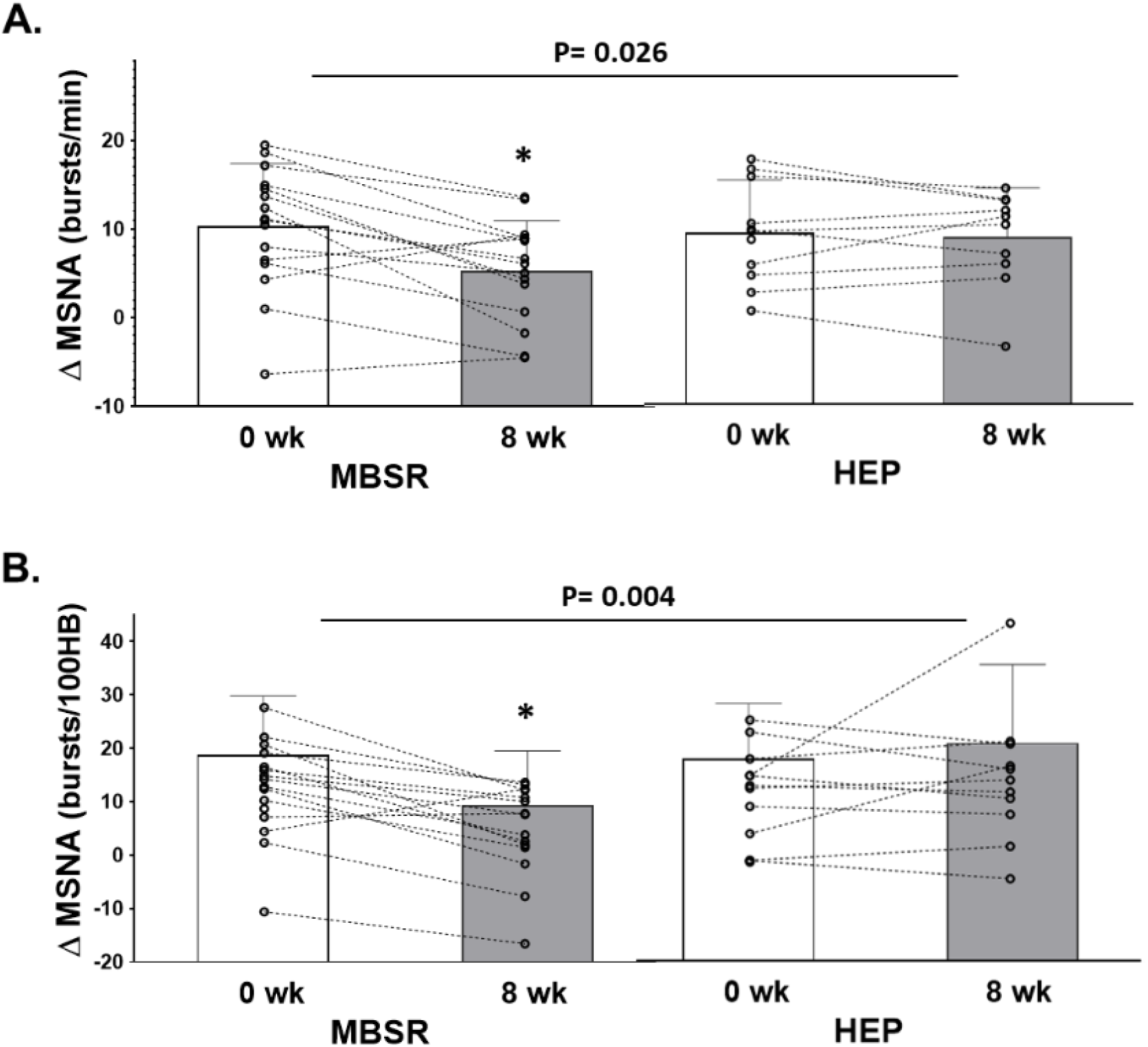
Reactivity of Muscle Sympathetic Nervous Activity (MSNA) Burst Frequency. (**A**) and MSNA Burst Incidence (**B**) averaged over 3 minutes of Mental Arithmetic from baseline (**0 wk**) to post-intervention (**8 wk**) in patients with chronic kidney disease randomized to Mindfulness-Based Stress Reduction (MBSR; N=16) versus Health Education Program (HEP; N=12) groups. Open circles (O) depict individual values for each study participant and bar graphs depict the mean and standard deviation values at baseline and post-intervention within each group. P-values denote statistically significant differences in the change from baseline to post-intervention between groups (Group x Time interaction) by repeated ANOVA tests. * denotes statistically significant within-group difference from baseline to post-intervention by paired *t*-tests.

**Figure 3.**
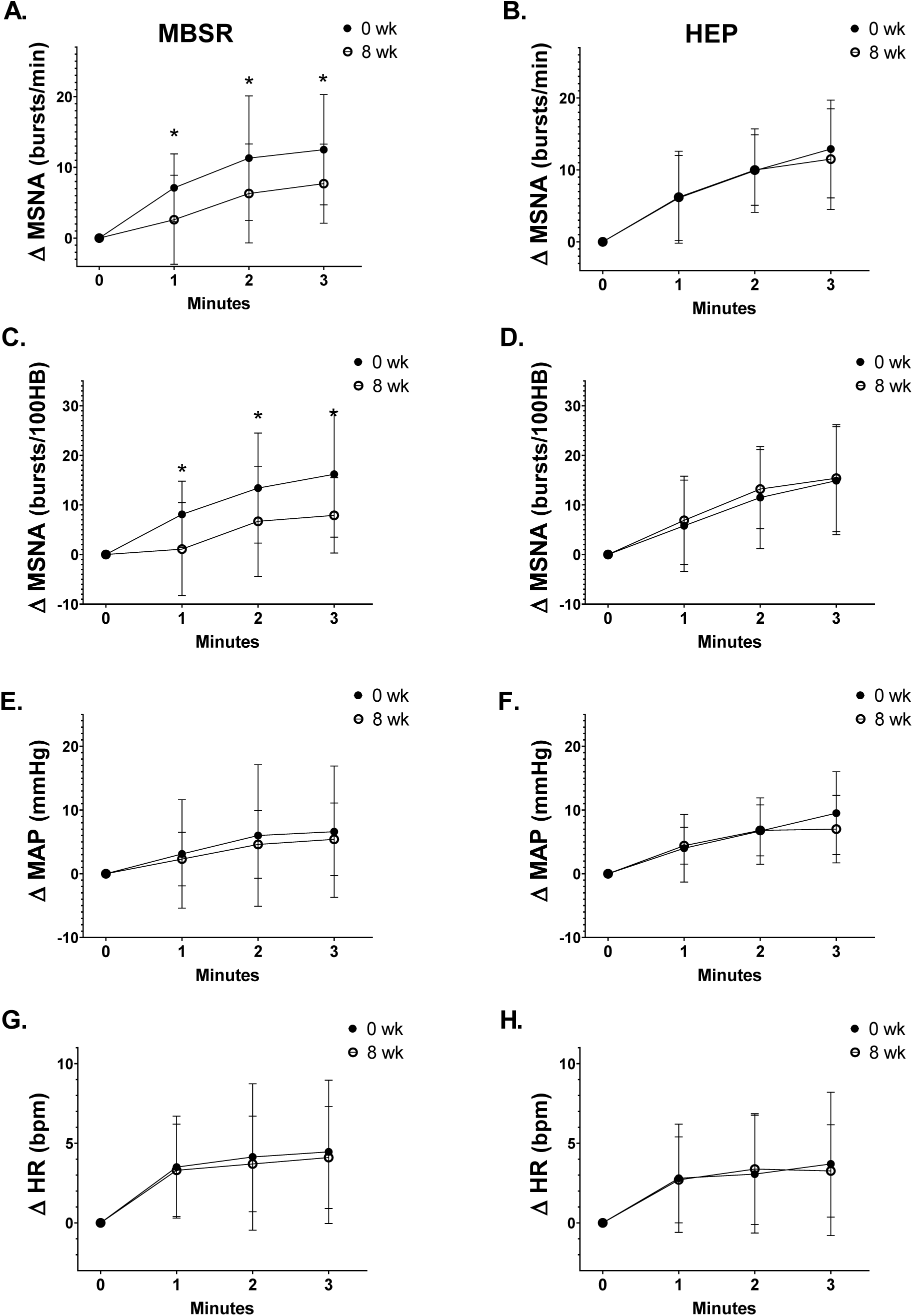
Reactivity in Muscle Sympathetic Nervous Activity (MSNA) Burst Frequency. (**A**, **B**), MSNA Burst Incidence (**C**, **D**), Mean Arterial Blood Pressure (MAP) (**E**, **F**) and Heart Rate (HR) (**G**, **H**) during each minute of Mental Arithmetic from baseline (**0 wk**, closed circles) to post-intervention (**8 wk**, open circles) in patients with chronic kidney disease randomized to Mindfulness-Based Stress Reduction (MBSR; N=16) versus Health Education Program (HEP; N=12) groups. * denotes a statistically significant Time (baseline vs. post-intervention) by Minute (1^st^, 2^nd^ or 3^rd^ minute) interaction effect within the MSBR group by repeated ANOVA tests.

#### Reactivity to Static Handgrip Exercise

The mean change in MSNA frequency, BP and HR over 3 minutes of SHG were not significantly changed from baseline to post-intervention in both MBSR and HEP groups (p>0.05 for both), although the MSNA burst incidence reactivity to SHG decreased within the MBSR group (p<0.05), but not within the HEP group (**Table 2**). LMM analyses showed no significant group difference in the slope-of-rise of absolute MSNA changes during 3 minutes of SHG from baseline to post-intervention between the MBSR and HEP groups (effect sizes for the group difference for MSNA: –0.59 bursts/min per min; p = 0.43 and –1.58 bursts/100 heartbeats per min; p = 0.14).

#### Reactivity to Cold Pressor Test

MSNA, BP and heart rate reactivity to CPT remained unchanged in both groups from baseline to post-intervention in both MBSR and HEP groups (**Table 2**). Perceived pain levels during CPT were comparable between baseline and post-intervention in both groups (2.3 ± 1.3 to 2.0 ± 1.4 in the MBSR and 3.5 ± 0.7 to 2.9 ± 1.0 in the HEP group, p=0.416 for Time by Group interaction).

### The Impact of Baseline MSNA Reactivity on MBSR-Induced Changes

We further examined the linear association of baseline levels of MSNA reactivity during mental arithmetic with MBSR-induced changes in MSNA reactivity within the MBSR group. The magnitude of change in MSNA reactivity was inversely associated with baseline MSNA reactivity (p<0.001, **Figure 4A, C**) in the MBSR group, while no significant association was found within the HEP group (p>0.05; **Figure 4B, D**).

**Figure 4.**
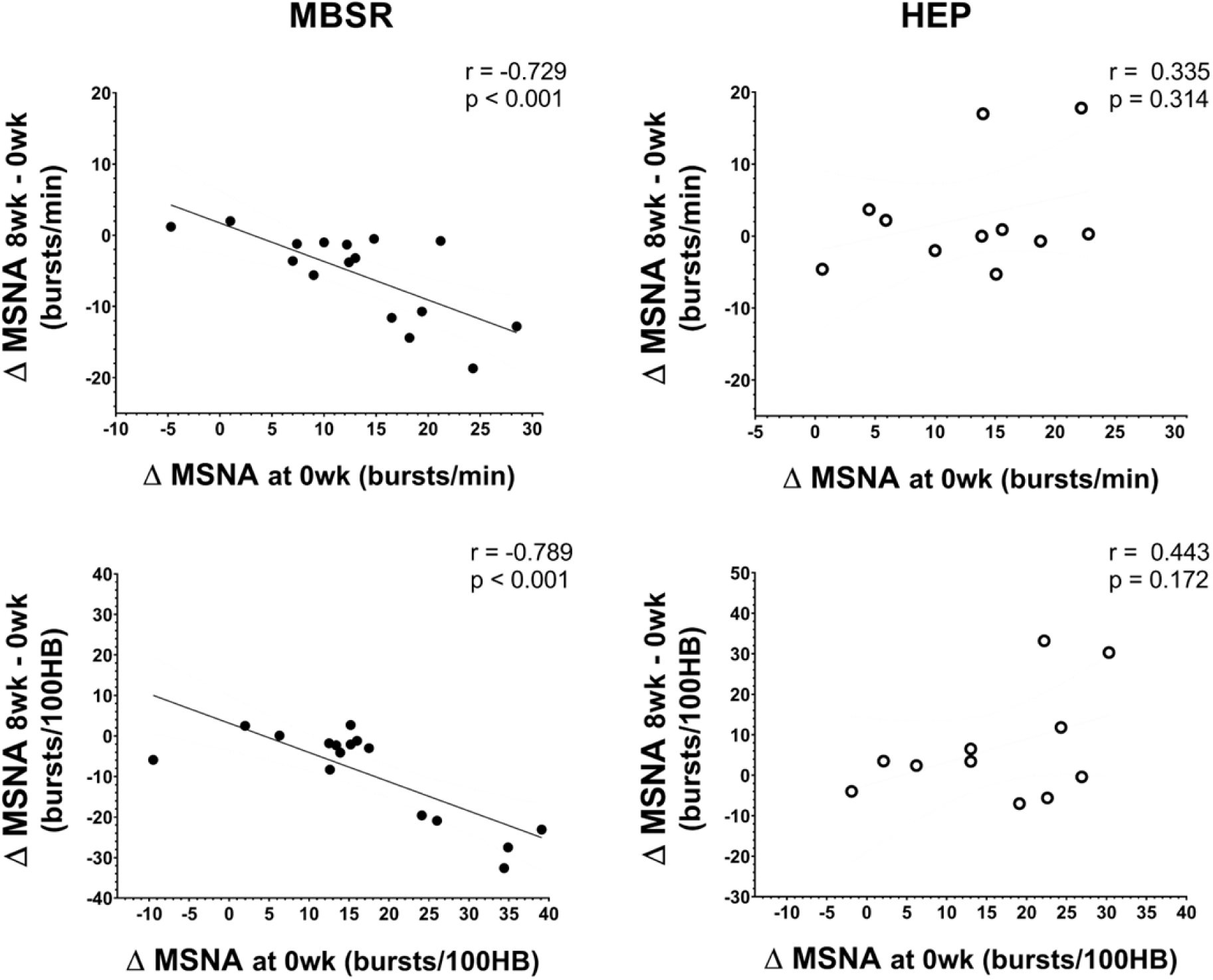
Association between Reactivity in Muscle Sympathetic Nervous activity (MSNA) Burst Frequency and MSNA Burst Incidence during mental arithmetic at Baseline (**0 wk**) and Change (post-intervention; **8 wk** – baseline; **0 wk**) in MBSR (**A, C** respectively, n=16) and in the HEP Group (**B, D** respectively, n=12) by Pearson correlation tests. Closed circles (•) depict individual values for each study participant in the MBSR group while open circles (O) depict individual values for each study participant in the HEP group.

## Discussion

We conducted a randomized controlled trial testing the efficacy of MBSR in lowering SNS overactivity in patients with CKD patients. By using direct intraneural recordings of SNS activity directed to the muscle via microneurography, we demonstrate a significant reduction in MSNA reactivity during acute mental stress in CKD patients who were randomized to 8 weeks of MBSR compared to the HEP control intervention. The MBSR intervention was well-accepted with high adherence rates to classes and practice homework, and our results suggest that MBSR may be a feasible adjunct intervention to improve CV risk by modulating autonomic balance in CKD.

MBSR is the most extensively studied meditation intervention in the US, with confirmed psychological and physiological health benefits including improved CV disease risk across different patient populations ^13–15, 19, 20^. Mindfulness-based interventions are hypothesized to improve CV health by promoting effective self-emotion regulation through non-judgmental present-moment awareness and a shift in stress appraisal, ultimately reducing the stress-reactivity response ^27, 28, 47^. These improvements in coping skills and psychological functioning may be coupled with the normalization of physiological functioning, such as the autonomic nervous system, potentially leading to improved CV outcomes ^29^. Despite this concept that autonomic regulation during stress may underlie the CV health benefits of MBSR, limited research has investigated this area, with existing data primarily relying on indirect markers of autonomic function ^30–32^ such as heart rate variability and plasma norepinephrine ^33^, and largely focusing on resting-state activity. We now provide the first evidence that MBSR may modulate SNS response to acute mental stress in CKD, a patient population at increased CV risk by virtue of chronic SNS overactivation. Our results align with previous evidence demonstrating that MBSR has favorable effects on stress responses on objectively measured stress biomarkers ^23–25^ as well as self-reported stress scales ^21, 22^. Prior work has shown that mindfulness-based interventions may attenuate increases in BP ^23, 24^, neurohormonal (cortisol, adrenocorticotropic hormone) and inflammatory (IL-6, TNF-a) reactivity ^25^, perceived stress levels ^21, 22^ and functional magnetic resonance imaging-based neural reactivity ^26, 48^ during stressful stimuli. Given heightened physiological responses to stressors have been linked to an increased risk of CV diseases and mortality ^49–51^, our results demonstrating MBSR-driven reductions in SNS stress reactivity has important clinical implication for CV risk management in CKD.

Our data also demonstrate that participants who initially displayed an augmented MSNA response to stress exhibited a more substantial reduction in MSNA reactivity following the MBSR intervention. This pattern of intervention response linked to the initial stress profile was specific to MBSR and not observed within the HEP group; thus, it is unlikely that these findings can be explained by regression towards the mean. These findings are in line with previous reports demonstrating a greater degree of BP reduction with MBSR in those with elevated levels of BP initially ^52, 53^. Therefore, our findings suggest that the expected benefits of MBSR on autonomic stress reactivity may vary depending on an individual’s initial stress response patterns, emphasizing the importance of tailored interventions to maximize the clinical utility of MBSR for optimal CV risk management in CKD.

Contrary to our hypothesis, decreased MSNA reactivity was not significantly associated with changes in BP reactivity, nor did resting BP and MSNA change significantly following the MBSR intervention in the present study. Many ^19, 20^, but not all meta-analyses ^54^, have demonstrated clinically significant reductions in BP as well as task-driven BP responses after participating in a MBSR intervention in individuals with CV disease. Our previous investigation showed BP and MSNA were acutely attenuated during a mindfulness meditation session that was sustained during the immediate recovery period in CKD patients ^34^. However, in the present study, the acute effects of mindfulness meditation on resting BP and MSNA were not maintained after completion of 8 weeks of MBSR, although the time-frames and the settings of measurements were not comparable (i.e., the acute responses to a 14 minutes of mindfulness meditation session vs. the changes over 8 weeks on resting MSNA and responses to a stressor). It is possible that more comprehensive or prolonged practice is needed for sustained improvements in resting MSNA, with downstream improvements in BP profiles in CKD. Alternatively, long-term change in BP stress reactivity may be more closely tied to peripheral vascular mechanisms (i.e., adrenergic receptor sensitivity) rather than changes in central SNS output, or due to other non-adrenergic mechanisms (i.e., inflammatory or renin-angiotensin aldosterone system (RAAS) pathways), which can be further investigated in future studies. It should also be noted that the majority of study participants were on anti-hypertensive medications including RAAS inhibitors (76%), calcium-channel blockers (57%) and beta-blockers (36%), which might have modulated the transduction of SNS activation on peripheral vasculature during an acute stressor to some extent.

While we observed reductions in MSNA reactivity after MBSR during specifically mental stress induced by mental arithmetic, we did not observe similar changes in MSNA reactivity during different stressors (physical stress by handgrip exercise and pain stress induced by cold pressor test). Although the mechanisms underlying these discrepant results are unclear, these differences in autonomic stress reactivity after MBSR may be attributed to the unique physiological mechanisms underlying each stressor. Mental stress increases SNS activity primarily through central neural pathways involving higher brain centers such as the prefrontal cortex, amygdala, and hippocampus ^55, 56^. Mindfulness practice aims to promote emotional self-regulation and reduce stress appraisal, potentially achieved through enhanced regulation of central neural signals. Previous evidence has demonstrated that mindfulness training can reduce CV responses during mental stress tasks ^23, 24^, and induce positive changes in the brain regions responsible for processing stress neural signals ^57–60^. Therefore, MBSR may have greater effects on SNS activation during mental stress as opposed to other types of stressors that involves a multifaceted interplay of peripheral and central mechanisms to activate SNS activity ^61–63^.

### Limitations

The majority of participants were Black males which may limit the generalizability of study results to females and other races. Antihypertensive medications such as angiotensin receptor blockers are known to impact sympathetic activity ^64^, and therefore, may have confounded results, although no difference in medication usage was found between groups at baseline, and the primary results remained the same when adjusted for medication use. Furthermore, participants maintained a stable medication regimen throughout the study period. Therefore, MBSR-induced changes in MSNA stress reactivity are likely to represent true intervention effects, and the results of this study can be applicable to the general medicated CKD patients. The intervention sessions were administered virtually due to COVID-19 restrictions. However, besides the virtual platform, MBSR was administered per standard protocol by a certified MBSR trainer. The nature of the interventions did not allow for masking of the MBSR and HEP instructors and participants. However, analyses of all physiologic data were made in a masked fashion without the investigators’ knowledge of group assignment. In addition, the active control intervention, HEP, was administered in parallel and matched for group setting, instructor attention, intervention duration, time spent on at-home practice while omitting any components of mindfulness.

## Conclusion

In this randomized controlled trial, we evaluated the effects of MBSR on SNS activity and reactivity in CKD and demonstrated a beneficial effect of MBSR on sympathetic reactivity during mental stress. We also observed greater reductions in MSNA reactivity in participants with higher MSNA reactivity at baseline suggesting that MBSR may have greater beneficial effects in patients with higher sympathetic reactivity to stress. Our results highlight the therapeutic potential of MBSR in ameliorating mental stress-induced SNS overactivation in CKD.

## Institution Where Work Was Performed

The Human Integrative Physiology Laboratory, Division of Renal Medicine, Emory University, Atlanta, Georgia, 30322, USA

## Data Availability

The data that support the findings of this study are not openly available due to reasons of sensitivity and are available from the corresponding author upon reasonable request.

Author Contributions

J.J. and J.P. conceived and designed research; J.J, M.Z., D.D. and J.P. performed experiments; J.J., Y.H., and J.P. analyzed data; J.J., Y.H., M.Z., D.D., S.L. and J.P. interpreted results of experiments; J.J., Y.H., and J.P. prepared figures; J.J., Y.H., M.Z. and J.P. drafted manuscript; J.J., Y.H., M.Z., D.D., S.L. and J.P. edited and revised manuscript; J.J., Y.H., M.Z., D.D., S.L. and J.P. approved final version of manuscript.

## References

1. Go AS, Chertow GM, Fan D, McCulloch CE and Hsu CY. Chronic kidney disease and the risks of death, cardiovascular events, and hospitalization. N Engl J Med. 2004;351:1296–305.

2. Herzog CA, Asinger RW, Berger AK, Charytan DM, Diez J, Hart RG, Eckardt KU, Kasiske BL, McCullough PA, Passman RS, DeLoach SS, Pun PH and Ritz E. Cardiovascular disease in chronic kidney disease. A clinical update from Kidney Disease: Improving Global Outcomes (KDIGO). Kidney Int. 2011;80:572–86.

3. Grassi G, Quarti-Trevano F, Seravalle G, Arenare F, Volpe M, Furiani S, Dell’Oro R and Mancia G. Early sympathetic activation in the initial clinical stages of chronic renal failure. Hypertension. 2011;57:846–51.

4. Klein IH, Ligtenberg G, Neumann J, Oey PL, Koomans HA and Blankestijn PJ. Sympathetic nerve activity is inappropriately increased in chronic renal disease. J Am Soc Nephrol. 2003;14:3239–44.

5. Neumann J, Ligtenberg G, Klein, II, Koomans HA and Blankestijn PJ. Sympathetic hyperactivity in chronic kidney disease: pathogenesis, clinical relevance, and treatment. Kidney Int. 2004;65:1568–76.

6. Badve SV, Roberts MA, Hawley CM, Cass A, Garg AX, Krum H, Tonkin A and Perkovic V. Effects of beta-adrenergic antagonists in patients with chronic kidney disease: a systematic review and meta-analysis. J Am Coll Cardiol. 2011;58:1152–61.

7. Tomiyama H and Yamashina A. Beta-Blockers in the Management of Hypertension and/or Chronic Kidney Disease. Int J Hypertens. 2014;2014:919256.

8. Bakris GL, Fonseca V, Katholi RE, McGill JB, Messerli FH, Phillips RA, Raskin P, Wright JT, Jr., Oakes R, Lukas MA, Anderson KM, Bell DS and Investigators G. Metabolic effects of carvedilol vs metoprolol in patients with type 2 diabetes mellitus and hypertension: a randomized controlled trial. JAMA. 2004;292:2227–36.

9. Hundemer GL, Knoll GA, Petrcich W, Hiremath S, Ruzicka M, Burns KD, Edwards C, Bugeja A, Rhodes E and Sood MM. Kidney, Cardiac, and Safety Outcomes Associated With alpha-Blockers in Patients With CKD: A Population-Based Cohort Study. Am J Kidney Dis. 2021;77:178–189 e1.

10. Vonend O, Marsalek P, Russ H, Wulkow R, Oberhauser V and Rump LC. Moxonidine treatment of hypertensive patients with advanced renal failure. J Hypertens. 2003;21:1709–17.

11. Whelton PK, Carey RM, Aronow WS, Casey DE, Jr., Collins KJ, Dennison Himmelfarb C, DePalma SM, Gidding S, Jamerson KA, Jones DW, MacLaughlin EJ, Muntner P, Ovbiagele B, Smith SC, Jr., Spencer CC, Stafford RS, Taler SJ, Thomas RJ, Williams KA, Sr., Williamson JD and Wright JT, Jr. 2017 ACC/AHA/AAPA/ABC/ACPM/AGS/APhA/ASH/ASPC/NMA/PCNA Guideline for the Prevention, Detection, Evaluation, and Management of High Blood Pressure in Adults: A Report of the American College of Cardiology/American Heart Association Task Force on Clinical Practice Guidelines. J Am Coll Cardiol. 2018;71:e127–e248.

12. James PA, Oparil S, Carter BL, Cushman WC, Dennison-Himmelfarb C, Handler J, Lackland DT, LeFevre ML, MacKenzie TD, Ogedegbe O, Smith SC, Jr., Svetkey LP, Taler SJ, Townsend RR, Wright JT, Jr., Narva AS and Ortiz E. 2014 evidence-based guideline for the management of high blood pressure in adults: report from the panel members appointed to the Eighth Joint National Committee (JNC 8). JAMA. 2014;311:507–20.

13. Lakhan SE and Schofield KL. Mindfulness-based therapies in the treatment of somatization disorders: a systematic review and meta-analysis. PLoS One. 2013;8:e71834.

14. Lengacher CA, Kip KE, Barta M, Post-White J, Jacobsen PB, Groer M, Lehman B, Moscoso MS, Kadel R, Le N, Loftus L, Stevens CA, Malafa MP and Shelton MM. A pilot study evaluating the effect of mindfulness-based stress reduction on psychological status, physical status, salivary cortisol, and interleukin-6 among advanced-stage cancer patients and their caregivers. J Holist Nurs. 2012;30:170–85.

15. Lengacher CA, Shelton MM, Reich RR, Barta MK, Johnson-Mallard V, Moscoso MS, Paterson C, Ramesar S, Budhrani P, Carranza I, Lucas J, Jacobsen PB, Goodman MJ and Kip KE. Mindfulness based stress reduction (MBSR(BC)) in breast cancer: evaluating fear of recurrence (FOR) as a mediator of psychological and physical symptoms in a randomized control trial (RCT). J Behav Med. 2014;37:185–95.

16. Barnes VA, Pendergrast RA, Harshfield GA and Treiber FA. Impact of breathing awareness meditation on ambulatory blood pressure and sodium handling in prehypertensive African American adolescents. Ethn Dis. 2008;18:1–5.

17. Goldstein CM, Josephson R, Xie S and Hughes JW. Current perspectives on the use of meditation to reduce blood pressure. Int J Hypertens. 2012;2012:578397.

18. Manikonda JP, Stork S, Togel S, Lobmuller A, Grunberg I, Bedel S, Schardt F, Angermann CE, Jahns R and Voelker W. Contemplative meditation reduces ambulatory blood pressure and stress-induced hypertension: a randomized pilot trial. J Hum Hypertens. 2008;22:138–40.

19. Younge JO, Gotink RA, Baena CP, Roos-Hesselink JW and Hunink MG. Mind-body practices for patients with cardiac disease: a systematic review and meta-analysis. Eur J Prev Cardiol. 2015;22:1385–98.

20. Scott-Sheldon LAJ, Gathright EC, Donahue ML, Balletto B, Feulner MM, DeCosta J, Cruess DG, Wing RR, Carey MP and Salmoirago-Blotcher E. Mindfulness-Based Interventions for Adults with Cardiovascular Disease: A Systematic Review and Meta-Analysis. Ann Behav Med. 2020;54:67–73.

21. Hoge EA, Bui E, Marques L, Metcalf CA, Morris LK, Robinaugh DJ, Worthington JJ, Pollack MH and Simon NM. Randomized controlled trial of mindfulness meditation for generalized anxiety disorder: effects on anxiety and stress reactivity. J Clin Psychiatry. 2013;74:786–92.

22. Creswell JD, Pacilio LE, Lindsay EK and Brown KW. Brief mindfulness meditation training alters psychological and neuroendocrine responses to social evaluative stress. Psychoneuroendocrinology. 2014;44:1–12.

23. Lindsay EK, Young S, Smyth JM, Brown KW and Creswell JD. Acceptance lowers stress reactivity: Dismantling mindfulness training in a randomized controlled trial. Psychoneuroendocrinology. 2018;87:63–73.

24. Nyklicek I, Mommersteeg PM, Van Beugen S, Ramakers C and Van Boxtel GJ. Mindfulness-based stress reduction and physiological activity during acute stress: a randomized controlled trial. Health Psychol. 2013;32:1110–3.

25. Hoge EA, Bui E, Palitz SA, Schwarz NR, Owens ME, Johnston JM, Pollack MH and Simon NM. The effect of mindfulness meditation training on biological acute stress responses in generalized anxiety disorder. Psychiatry Res. 2018;262:328–332.

26. Dutcher JM, Boyle CC, Eisenberger NI, Cole SW and Bower JE. Neural responses to threat and reward and changes in inflammation following a mindfulness intervention. Psychoneuroendocrinology. 2021;125:105114.

27. Shapiro SL, Carlson LE, Astin JA and Freedman B. Mechanisms of mindfulness. J Clin Psychol. 2006;62:373–86.

28. Loucks EB, Schuman-Olivier Z, Britton WB, Fresco DM, Desbordes G, Brewer JA and Fulwiler C. Mindfulness and Cardiovascular Disease Risk: State of the Evidence, Plausible Mechanisms, and Theoretical Framework. Curr Cardiol Rep. 2015;17:112.

29. Creswell JD and Lindsay EK. How Does Mindfulness Training Affect Health? A Mindfulness Stress Buffering Account. Current Directions in Psychological Science. 2014;23:401–407.

30. Ditto B, Eclache M and Goldman N. Short-term autonomic and cardiovascular effects of mindfulness body scan meditation. Ann Behav Med. 2006;32:227–34.

31. Nijjar PS, Puppala VK, Dickinson O, Duval S, Duprez D, Kreitzer MJ and Benditt DG. Modulation of the autonomic nervous system assessed through heart rate variability by a mindfulness based stress reduction program. Int J Cardiol. 2014;177:557–9.

32. Wang SJ, Chang YC, Hu WY, Chang YM and Lo C. The Comparative Effect of Reduced Mindfulness-Based Stress on Heart Rate Variability among Patients with Breast Cancer. Int J Environ Res Public Health. 2022;19.

33. Curiati JA, Bocchi E, Freire JO, Arantes AC, Braga M, Garcia Y, Guimaraes G and Fo WJ. Meditation reduces sympathetic activation and improves the quality of life in elderly patients with optimally treated heart failure: a prospective randomized study. J Altern Complement Med. 2005;11:465–72.

34. Park J, Lyles RH and Bauer-Wu S. Mindfulness meditation lowers muscle sympathetic nerve activity and blood pressure in African-American males with chronic kidney disease. Am J Physiol Regul Integr Comp Physiol. 2014;307:R93–R101.

35. Levey AS, Coresh J, Greene T, Stevens LA, Zhang YL, Hendriksen S, Kusek JW, Van Lente F and Chronic Kidney Disease Epidemiology C. Using standardized serum creatinine values in the modification of diet in renal disease study equation for estimating glomerular filtration rate. Ann Intern Med. 2006;145:247–54.

36. Eisendrath SJ, Gillung E, Delucchi KL, Segal ZV, Nelson JC, McInnes LA, Mathalon DH and Feldman MD. A Randomized Controlled Trial of Mindfulness-Based Cognitive Therapy for Treatment-Resistant Depression. Psychother Psychosom. 2016;85:99–110.

37. Eisendrath SJ, Gillung EP, Delucchi KL, Chartier M, Mathalon DH, Sullivan JC, Segal ZV and Feldman MD. Mindfulness-based cognitive therapy (MBCT) versus the health-enhancement program (HEP) for adults with treatment-resistant depression: a randomized control trial study protocol. BMC Complement Altern Med. 2014;14:95.

38. MacCoon DG, Imel ZE, Rosenkranz MA, Sheftel JG, Weng HY, Sullivan JC, Bonus KA, Stoney CM, Salomons TV, Davidson RJ and Lutz A. The validation of an active control intervention for Mindfulness Based Stress Reduction (MBSR). Behav Res Ther. 2012;50:3–12.

39. Wallin BG and Fagius J. Peripheral sympathetic neural activity in conscious humans. Annu Rev Physiol. 1988;50:565–76.

40. Jeleazcov C, Krajinovic L, Munster T, Birkholz T, Fried R, Schuttler J and Fechner J. Precision and accuracy of a new device (CNAPTM) for continuous non-invasive arterial pressure monitoring: assessment during general anaesthesia. Br J Anaesth. 2010;105:264–72.

41. Mano T, Iwase S and Toma S. Microneurography as a tool in clinical neurophysiology to investigate peripheral neural traffic in humans. Clin Neurophysiol. 2006;117:2357–84.

42. Attkisson CC and Zwick R. The client satisfaction questionnaire. Psychometric properties and correlations with service utilization and psychotherapy outcome. Eval Program Plann. 1982;5:233–7.

43. Fonkoue IT, Hu Y, Jones T, Vemulapalli M, Sprick JD, Rothbaum B and Park J. Eight weeks of device-guided slow breathing decreases sympathetic nervous reactivity to stress in posttraumatic stress disorder. Am J Physiol Regul Integr Comp Physiol. 2020;319:R466–R475.

44. Park J, Marvar PJ, Liao P, Kankam ML, Norrholm SD, Downey RM, McCullough SA, Le NA and Rothbaum BO. Baroreflex dysfunction and augmented sympathetic nerve responses during mental stress in veterans with post-traumatic stress disorder. J Physiol. 2017;595:4893–4908.

45. Mark AL, Victor RG, Nerhed C and Wallin BG. Microneurographic studies of the mechanisms of sympathetic nerve responses to static exercise in humans. Circ Res. 1985;57:461–9.

46. Victor RG, Leimbach WN, Jr., Seals DR, Wallin BG and Mark AL. Effects of the cold pressor test on muscle sympathetic nerve activity in humans. Hypertension. 1987;9:429–36.

47. Garland EL, Farb NA, Goldin P and Fredrickson BL. Mindfulness Broadens Awareness and Builds Eudaimonic Meaning: A Process Model of Mindful Positive Emotion Regulation. Psychol Inq. 2015;26:293–314.

48. Goldin PR and Gross JJ. Effects of mindfulness-based stress reduction (MBSR) on emotion regulation in social anxiety disorder. Emotion. 2010;10:83–91.

49. Schultz MG, Otahal P, Cleland VJ, Blizzard L, Marwick TH and Sharman JE. Exercise-induced hypertension, cardiovascular events, and mortality in patients undergoing exercise stress testing: a systematic review and meta-analysis. Am J Hypertens. 2013;26:357–66.

50. Weiss SA, Blumenthal RS, Sharrett AR, Redberg RF and Mora S. Exercise blood pressure and future cardiovascular death in asymptomatic individuals. Circulation. 2010;121:2109–16.

51. Chida Y and Steptoe A. Greater cardiovascular responses to laboratory mental stress are associated with poor subsequent cardiovascular risk status: a meta-analysis of prospective evidence. Hypertension. 2010;55:1026–32.

52. Loucks EB, Nardi WR, Gutman R, Kronish IM, Saadeh FB, Li Y, Wentz AE, Webb J, Vago DR, Harrison A and Britton WB. Mindfulness-Based Blood Pressure Reduction (MB-BP): Stage 1 single-arm clinical trial. PLoS One. 2019;14:e0223095.

53. Lee EKP, Yeung NCY, Xu Z, Zhang D, Yu CP and Wong SYS. Effect and Acceptability of Mindfulness-Based Stress Reduction Program on Patients With Elevated Blood Pressure or Hypertension: A Meta-Analysis of Randomized Controlled Trials. Hypertension. 2020;76:1992–2001.

54. Abbott RA, Whear R, Rodgers LR, Bethel A, Thompson Coon J, Kuyken W, Stein K and Dickens C. Effectiveness of mindfulness-based stress reduction and mindfulness based cognitive therapy in vascular disease: A systematic review and meta-analysis of randomised controlled trials. J Psychosom Res. 2014;76:341–51.

55. Ulrich-Lai YM and Herman JP. Neural regulation of endocrine and autonomic stress responses. Nat Rev Neurosci. 2009;10:397–409.

56. Herman JP, Figueiredo H, Mueller NK, Ulrich-Lai Y, Ostrander MM, Choi DC and Cullinan WE. Central mechanisms of stress integration: hierarchical circuitry controlling hypothalamo-pituitary-adrenocortical responsiveness. Front Neuroendocrinol. 2003;24:151–80.

57. Creswell JD, Way BM, Eisenberger NI and Lieberman MD. Neural correlates of dispositional mindfulness during affect labeling. Psychosom Med. 2007;69:560–5.

58. Holzel BK, Hoge EA, Greve DN, Gard T, Creswell JD, Brown KW, Barrett LF, Schwartz C, Vaitl D and Lazar SW. Neural mechanisms of symptom improvements in generalized anxiety disorder following mindfulness training. Neuroimage Clin. 2013;2:448–58.

59. Modinos G, Ormel J and Aleman A. Individual differences in dispositional mindfulness and brain activity involved in reappraisal of emotion. Soc Cogn Affect Neurosci. 2010;5:369–77.

60. Taren AA, Gianaros PJ, Greco CM, Lindsay EK, Fairgrieve A, Brown KW, Rosen RK, Ferris JL, Julson E, Marsland AL, Bursley JK, Ramsburg J and Creswell JD. Mindfulness meditation training alters stress-related amygdala resting state functional connectivity: a randomized controlled trial. Soc Cogn Affect Neurosci. 2015;10:1758–68.

61. Gallagher KM, Fadel PJ, Smith SA, Stromstad M, Ide K, Secher NH and Raven PB. The interaction of central command and the exercise pressor reflex in mediating baroreflex resetting during exercise in humans. Exp Physiol. 2006;91:79–87.

62. Burton AR, Fazalbhoy A and Macefield VG. Sympathetic Responses to Noxious Stimulation of Muscle and Skin. Front Neurol. 2016;7:109.

63. Smith SA, Mitchell JH and Garry MG. The mammalian exercise pressor reflex in health and disease. Exp Physiol. 2006;91:89–102.

64. Ohlstein EH, Brooks DP, Feuerstein GZ and Ruffolo RR, Jr. Inhibition of sympathetic outflow by the angiotensin II receptor antagonist, eprosartan, but not by losartan, valsartan or irbesartan: relationship to differences in prejunctional angiotensin II receptor blockade. Pharmacology. 1997;55:244–51.

